# LARGE CORE THROMBECTOMY: FEASIBILITY OF SIMPLIFIED PROTOCOL IN RESOURCE-LIMITED SETTINGS

**DOI:** 10.1101/2025.03.03.25323288

**Authors:** Thien Quang Le, Son Van Dang Nguyen, Tao Van Tran, Tuan Phuoc Pham, Nam Van Le, Dzung Thi Nguyen, Hoang Huy Nguyen, Hang Vu Nhat Pham, Toan Khac Ngo, Trung Quoc Nguyen, Thong Nhu Pham, Hieu Van Cao, Vu Thanh Huynh, Hai Quang Duong, Chih-Hao Chen, Trung Thanh Nguyen

**Author notes:** **Corresponding author:** Son Van Dang Nguyen, MD, MSc, Department of Stroke, Da Nang Hospital, Da Nang, Vietnam.

## Abstract

**Introduction:** Several trials have demonstrated the benefits of endovascular thrombectomy (EVT) for large-core strokes (ASPECTS < 6). However, its effectiveness in lower-middle-income countries with resource-limited settings remains uncertain. This study evaluated the feasibility of EVT for large-core strokes using a simplified imaging protocol with non-contrast CT (NCCT) and CT angiography (CTA) in a resource-constrained environment.

**Methods:** We conducted a prospective, single-center, observational study from May 2023 to May 2024 at Da Nang Hospital, Vietnam. Patients with anterior circulation large-vessel occlusion strokes, ASPECTS < 6 on NCCT, admission NIHSS ≥ 6, and EVT within 24 hours were included. The primary outcome was the modified Rankin Scale (mRS) score at 90 days. Functional independence was defined as mRS 0–2 and ambulatory independence as mRS 0–3. Safety outcomes included symptomatic intracranial hemorrhage (sICH). Outcomes were compared based on reperfusion success (mTICI ≥2b vs. 0–2a), ASPECTS (0–2 vs. 3–5), and time window (≤6 vs. >6 hours).

**Results:** Among 157 EVT-treated patients, 52 (33.1%) had ASPECTS < 6. The median age was 62.5 years, and 57.7% were male. Median onset-to-hospital time was 4.1 hours (IQR 1.8-7.9), admission NIHSS 15 (IQR 13-19.5), and initial ASPECTS 4 (IQR 3-4). Successful reperfusion (mTICI ≥2b) was achieved in 78.9%. At 90 days, the median mRS was 3.5 (IQR 3-5.5).

Functional independence was observed in 23.1% and ambulatory independence in 50%. sICH occurred in 9.6%, and mortality was 25%. Successful reperfusion was the only independent predictor of ambulatory independence (OR 14.7, 95% CI 1.6–134). Patients with ASPECTS 3–5 had significantly higher ambulatory independence than those with ASPECTS 0–2 (58.5% vs. 18.2%, p=0.017). No significant differences were found between early and late-window groups.

**Conclusion:** EVT is feasible for large-core stroke patients in lower-income countries using a simplified NCCT-CTA protocol. Successful reperfusion is a key determinant of improved outcomes.

## INTRODUCTION

Endovascular thrombectomy (EVT) is the gold standard for treating selected patients with acute ischemic stroke (AIS) caused by large vessel occlusions (LVO). Current guidelines recommend the highest level of evidence for EVT in patients with acute large vessel occlusions who have a small to medium infarct core, defined by an Alberta Stroke Program Early Computed Tomography Score (ASPECTS) of 6 or higher within a 24-hour window.^1^ Patients with acute large infarct cores are not typically candidates for endovascular treatment, although they account for approximately 25%.^2^ In recent years, the benefits of EVT for acute large infarct core (LIC) stroke have been demonstrated in high-quality trials^3-8^. However, these trials have only been conducted exclusively in high- or upper-middle-income countries and have primarily relied on magnetic resonance imaging (MRI) or perfusion imaging for patients selection.

In practical clinical scenarios, particularly in lower-income countries with limited resources, advanced imaging modalities are not widely available in emergency situations. Non-contrast computed tomography (NCCT) - computed tomography angiography (CTA) is still the most practical and accessible imaging approach for stroke evaluation. Despite their widespread use, the application of NCCT-based selection for EVT in large-core infarct patients remains a subject of ongoing debate. Additionally, in these countries, comprehensive stroke centers are often overwhelmed, along with a lack of both experience and robust evidence on the outcomes of large-core thrombectomy, making its implementation in clinical practice cautious. Consequently, the efficacy of large-core thrombectomy has not been sufficiently proven and continues to be a subject of uncertainty in lower-income countries.

NCCT is a simple, rapid, and readily available imaging modality for assessing early ischemic changes, making it a key part of stroke assessment in most centers worldwide. The TENSION trial demonstrated that NCCT-based selection for EVT in patients with ASPECTS scores of 3–5 was effective within a 12-hour window.^6^ Similarly, while the TESLA trial yielded negative results for patients with ASPECTS of 2–5 in the 24-hour window, it still suggests a potential role for thrombectomy using NCCT-based selection within this timeframe.^8^

Given the widespread availability of NCCT and its practicality in resource-limited settings, its role in guiding EVT decisions for patients with large infarct cores warrants further investigation. In this single-center, real-world observational study conducted in Vietnam - a lower-middle-income country, we aimed to evaluate the efficacy and safety of EVT within 24 hours in patients with non-restrictive acute large-core stroke (ASPECTS < 6) based on NCCT selection.

## METHODS

### Study design and eligible patients

This study was designed as a prospective, single-center, observational study over a one-year period from May 2023 to May 2024, conducted at Da Nang Hospital, Vietnam. Inclusion criteria were: age ≥ 18; acute ischemic stroke due to LVO in the anterior circulation, confirmed by CTA showing occlusion of the internal carotid artery (ICA) or the M1 segment of the middle cerebral artery (MCA), or tandem occlusion; large infarct core, defined as an Alberta Stroke Program Early Computed Tomography Score (ASPECTS) < 6 on NCCT at admission; moderate to severe neurological deficit, with an admission National Institutes of Health Stroke Scale (NIHSS) ≥ 6; underwent EVT within 24 hours from symptom onset. Exclusion criteria were: pre-stroke disability (mRS ≥ 3); multiple or bilateral ischemic strokes; severe or end-stage medical conditions; severe coagulation disorders, and inability to provide consent or follow-up.

The primary efficacy outcome was evaluated by modified Rankin Scale (mRS) at 90 days post-EVT. Functional independence was defined as an mRS of 0–2, while ambulatory independence was described as an mRS of 0–3. Safety outcomes included symptomatic intracranial hemorrhage (sICH), as per the Safe Implementation of Thrombolysis in Stroke-Monitoring Study (SITS-MOST) criteria^9^ and the overall mortality within 90 days.

ASPECTS was initially assessed by two board-certified neurologists and subsequently confirmed by an experienced radiologist in case of disagreement. Early neurological deterioration (END) was defined as an increase in the NIHSS score by ≥ 4 points within 24 hours after thrombectomy. Collateral score was evaluated using the Tan et al. score (0-4), which classifies collaterals as ‘good’ if they are seen in ≥ 50% of the middle cerebral artery territory based on CTA.^10^

A two-step imaging protocol using NCCT and CT angiography (CTA) was applied for patient selection. No MRI or perfusion imaging was required. Recombinant tissue plasminogen activator (rt-PA) was considered for patients presenting within the 4.5-hour time window, with administration determined at the discretion of the treating physician. EVT techniques were performed at the discretion of the neurointerventionist. The degree of reperfusion was assessed using the modified Thrombolysis in Cerebral Infarction (mTICI) scale, with successful reperfusion defined as mTICI ≥ 2b.^11^ The 90-day follow-up mRS assessment was conducted via telephone or during outpatient visits.

This study was approved by the Ethics Committee of Da Nang Hospital (IRB No. 2449/BVDN-HÐYÐ). Informed consent was obtained from patients or their family members before inclusion in the study.

### Statistical analysis

All statistical analyses were conducted using Stata 17 (StataCorp LLC, College Station, TX, USA). Continuous variables were expressed as medians with interquartile ranges (IQRs), while categorical variables were presented as frequencies and percentages. Group comparisons were performed using the Chi-square test or Fisher’s exact test for categorical variables and the Mann-Whitney U test or independent T-test for continuous variables, depending on data distribution. To determine independent predictors of favorable outcomes (modified Rankin Scale 0-3 at 90 days), multivariable logistic regression analysis was conducted, adjusting for potential confounders. Patient characteristics were analyzed and compared across time windows (<6 hours vs. >6 hours), ASPECTS (0–2 vs. 3–5), and mTICI (0–2a vs. 2b–3) to identify differences between subgroups. Statistical significance was set at p < 0.05.

## RESULTS

### Patients characteristics

From May 2023 to May 2024, we screened 157 patients with acute ischemic stroke due to large vessel occlusions who underwent EVT within 24 hours of stroke onset. Of these, 52 patients (33.1%) had large infarct cores with an ASPECTS of less than 6 and met the study criteria. The 90-day follow-up was completed in August 2024, with no missing data for the primary outcome. The median age was 62.5 years (IQR 58.5-71), and 57.7% (30/52) of patients were men. The median NIHSS at admission was 15 (IQR 13-19.75), and the median ASPECTS on NCCT before the procedure was 4 (IQR 3-4) **(Table 1)**.

**Table 1.** Baseline characteristics.

| **Characteristics of the patients** | **N = 52** |
| --- | --- |
| **Age, years - Median (IQR)** | 62.5 (58.5-71.0) |
| **Male Sex - no.(%)** | 30 (57.7%) |
| **Pre-mRS - Median (IQR)** | 0 (0-0) |
| **History - no.(%)** |  |
| TIA/Stroke | 7 (13.5%) |
| Hypertension | 42 (80.8%) |
| Atrial Fibrillation | 16 (30.8%) |
| Diabetes mellitus | 6 (11.5%) |
| Anticoagulant | 8 (15.4%) |
| Antiplatelet | 10 (19.2%) |
| **Balines NIHSS - Median (IQR)** | 15.0 (13-19) |
| **Systolic blood pressure -Median (IQR)** | 140 (120-150) |
| **Balines ASPECTS - Median (IQR)** | 4 (3-4) |
| 0-2 - no.(%) | 11 (21.2%) |
| 3 - no.(%) | 14 (26.9%) |
| 4 - no.(%) | 15 (28.8%) |
| 5 - no.(%) | 12 (23.1%) |
| **Intravenous thrombolysis - no.(%)** | 16 (30.8%) |
| **Occlusion site - no.(%)** |  |
| ICA | 5 (9.6%) |
| M1 segment of MCA | 33 (63.5%) |
| Tandem | 14 (26.9%) |
| **Collateral score - Median (IQR)** | 2 (1-2) |
| **Stroke Etiology- no.(%)** |  |
| Large-Artery Atherosclerosis | 26 (50%) |
| Cardioembolic | 18 (34.6%) |
| Undetermined or other | 8 (15.4%) |
| **Median time from onset to admission (IQR) (hours)** | 4.1 (1.8-7.9) |
| ≤ 6 hours - no.(%) | 35 (67.3%) |
| 6-24 hours - no.(%) | 17 (32.7%) |
| **Median duration (IQR) (min)** |  |
| Door to Needle time | 40 (35-50) |
| Door to puncture time | 111 (83.0-134.5) |
| Door to reperfusion | 146.5 (124-185.5) |

Abbreviations: mRS (modified Rankin scale), TIA (Transient ischemic attack), NIHSS (National Institutes of Health Stroke Scale), ICA (Internal carotid artery), MCA (Middle cerebral artery).

Regarding ASPECTS distribution, 11 patients (21.2%) had ultra-large infarct cores (ASPECTS 0-2), while 41 patients (78.8%) had ASPECTS 3-5. The median time from the last known well to hospital admission was 4.1 hours (IQR 1.8-7.9), with 32.7% (17/52) of patients presenting in the late window (beyond 6 hours from stroke onset).

Intravenous thrombolysis was used as a bridging therapy in 30.8% (16/52) of patients. Most patients had occlusions in the M1 branch of the middle cerebral artery with 63.5% (33/52), while internal carotid artery occlusions occurred in 9.6% (5/52), and tandem lesions in 26.9% (14/52). The median collateral score was 1 (IQR 1-2), with 44.2% (23/52) of patients having good collaterals. The median number of passes was 2 (IQR 1-3.5). Successful recanalization (mTICI ≥ 2b) was achieved in 78.8% (41/52) of patients. The median door-to-puncture time was 110 minutes (IQR 83-134.5), and the median door-to-reperfusion time was 146.5 minutes (IQR 124-185.5).

### Outcomes

At 90 days, the median mRS was 3.5 (IQR 3-5.5) **(Figure 1)**. Functional independence (mRS 0-2) was achieved by 23.1% (12/52) of patients, while 50% (26/52) achieved an ambulatory outcome (mRS 0–3) **(Table 2)**. END occurred in 20 patients (38.5%). Decompressive craniectomy was performed in 5 patients (9.6%) within 7 days.

**Table 2.** Efficacy and Safety Outcomes and Treatment Effects.

| Outcomes | N = 52 |
| --- | --- |
| **Functional outcome** |  |
| Median mRS at 90 day (IQR) | 3.5 (3-5.5) |
| mRS 0-2 at 90 day - no. (%) | 12 (23.1%) |
| mRS 0-3 at 90 day - no. (%) | 26 (50%) |
| **Safety outcomes** |  |
| Symptomatic intracranial hemorrhage - no.(%) | 5 (9.6%) |
| Any intracranial hemorrhage - no.(%) | 31 (59.6%) |
| Death within 90 days- no.(%) | 13 (25%) |
| Early neurological deterioration - no.(%) | 20 (38.5%) |
| Decompressive hemicraniectomy - no.(%) | 5 (9.6%) |
| **Treatment Effects** |  |
| Final mTICI - no.(%) |  |
| 0-2a | 11 (21.2%) |
| 2b-3 | 41 (78.8%) |
| Median Pass of EVT (IQR) | 2 (1.0-3.5) |
| Median NIHSS at 24 hours (IQR) | 17.5 (12.0-24.0) |
| Median NIHSS at 7 days or discharge (IQR) | 15.0 (9.0-31.0) |

Abbreviations: mRS (modified Rankin scale), mTICI (The modified treatment in cerebral infarction), NIHSS (National Institutes of Health Stroke Scale), EVT (Endovascular Thrombectomy)

**Figure 1.**
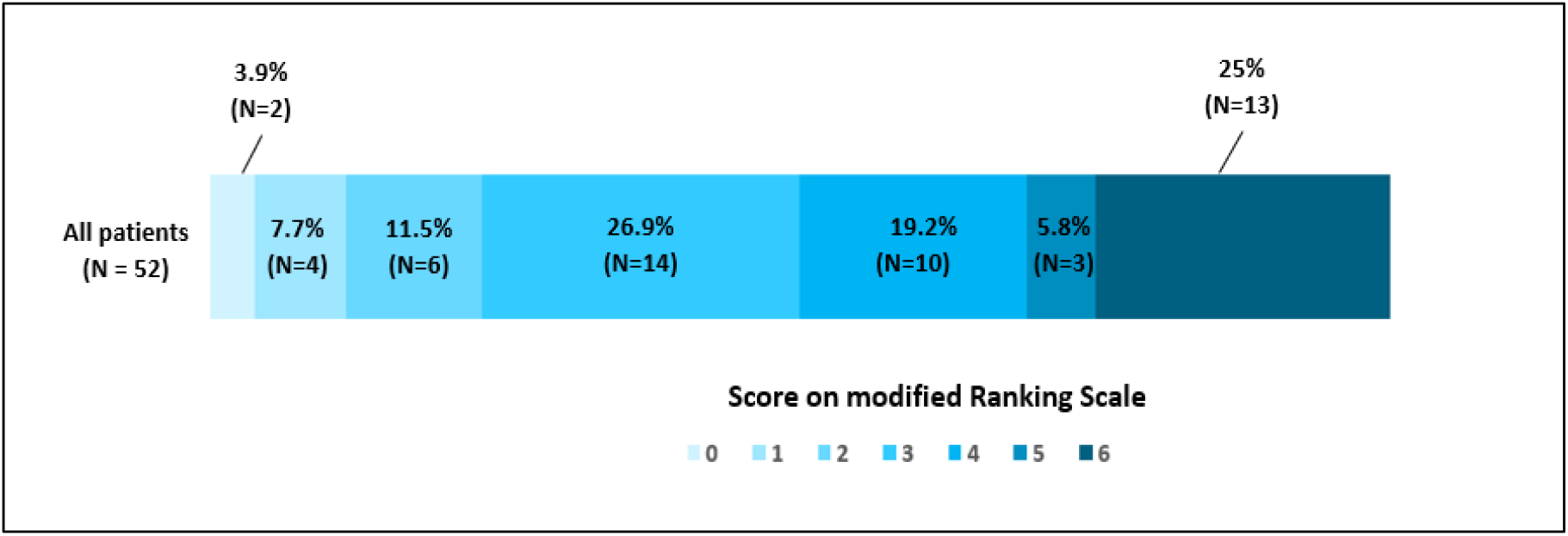
Distribution of mRS scores at 90 days.

Regarding safety outcomes, any intracerebral hemorrhage within 24 hours after EVT occurred in 59.6% (31/52) of patients, and symptomatic intracerebral hemorrhage was observed in 5 patients (9.6%). The mortality rate was 25% (13/52).

Multivariable logistic regression analysis identified successful recanalization as the only factor independently associated with ambulatory independence (mRS 0-3) (OR 14.7, 95% CI 1.6-134), after adjusting for age, admission NIHSS, and door-to-groin time.

A significant difference was observed in mRS at 90 days between the successful group (median 3, IQR 2-4) and unsuccessful recanalization (median 6, IQR 5-6, p=0.0008). The rate of ambulatory independence was 61% (25/41) versus 9.1% (1/11) respectively (p=0.005) **(Figure 2)**. The mortality rate was significantly lower in the successful recanalization group 17.1% (7/41) compared to the unsuccessful recanalization group 54.6% (6/11), with p=0.019. Although any intracerebral hemorrhage and symptomatic intracerebral hemorrhage occurred more frequently in the successful recanalization group, the difference was not statistically significant.

**Figure 2.**
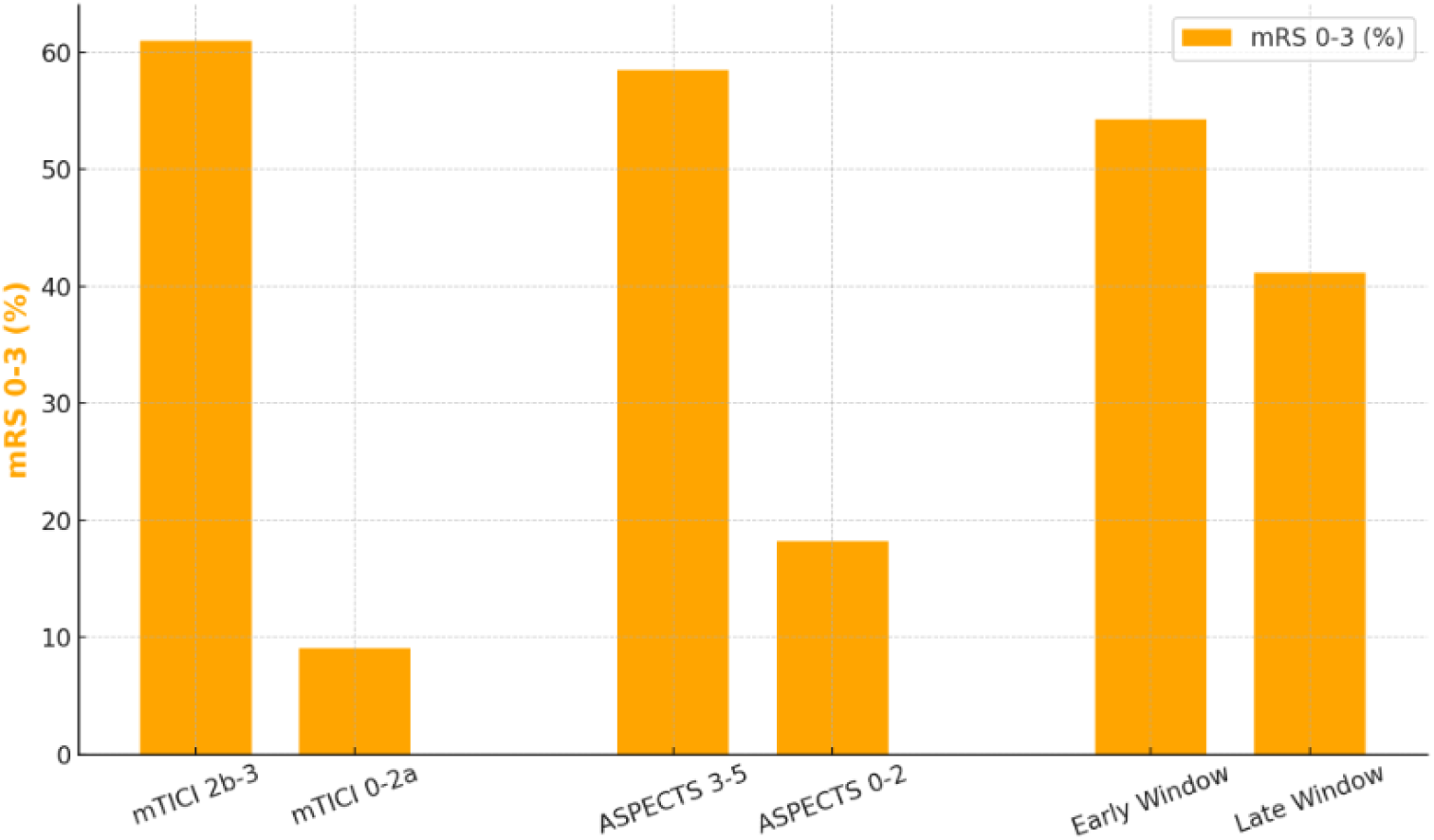
Comparison of Ambulatory Independence Across Subgroups. Abbreviations: mRS (modified Rankin scale), (ASPECTS) Alberta Stroke Program Early Computed Tomography Score

In the ASPECTS 3-5 group, 58.5% (24/41) of patients achieved an ambulatory outcome, whereas only 18.2% (2/11) did so in the ASPECTS 0-2 group (p=0.017) **(Figure 2)**. The ASPECTS 0-2 group also had higher rates of death, symptomatic intracerebral hemorrhage, and any intracerebral hemorrhage compared to the ASPECTS 3-5 group. We found no significant differences between the early (≤ 6 hours) and late window (> 6 hours) groups in an ambulatory outcome [54.3% (19/35) vs 41.2% (7/17), respectively, p=0.375] as well as other safety outcomes **(Figure 2)**.

## DISCUSSION

Our study provides real-world evidence that EVT remains effective and safe for LIC patients selected using NCCT-CTA alone, supporting the feasibility of simplified imaging protocols in resource-limited settings.

EVT has been confirmed to be effective and safe for patients with LICs within the 24-hour time window through large clinical trials conducted in upper-middle and high-income countries, where comprehensive stroke centers are well-equipped with timely resources and operate under well-managed overload conditions. A meta-analysis of large clinical trials found that EVT significantly increased the likelihood of achieving mRS 0-2 at 90 days (19.5% vs. 7.5%, RR 2.49 [95% CI, 1.92-3.24]), further validating EVT’s role in LIC patients.^12^ NCCT is the most commonly available stroke imaging modality worldwide, and its speed, accessibility, and cost-effectiveness make it a valuable tool for EVT decision-making. NCCT is not only applicable within the standard 6-hour window but has also been proven non-inferior to advanced imaging and is broadly accepted for selecting patients with large vessel occlusion and non-large infarct cores in the extended 6-to 24-hour window, making it a viable alternative in centers with limited access.^13,14^ In our study, NCCT was used to assess large infarct core volume with ASPECTS < 6, and CTA confirmed vessel occlusion, eliminating the need for advanced imaging. The feasibility of this approach is essential for expanding EVT eligibility in lower-resource stroke centers, where MRI and CTP are often unavailable or impractical due to cost, infrastructure limitations, and prolonged acquisition times in an already overwhelmed emergency setting. The TENSION trial, which exclusively used NCCT-based ASPECTS (3-5) selection, confirmed that EVT is beneficial in LIC patients without the need for MRI or perfusion imaging.^6^ When comparing our results with other large RCTs **(Table 3)**, the functional independence rate (mRS 0-2 at 90 days) in large-core trials has ranged from 13.3% to 30%, and our study’s 23.1% functional independence rate aligns with these findings. The results in this study were particularly comparable to the ANGEL-ASPECTS trial in terms of ambulatory outcomes and were better than other trials, with 50% of patients achieving mRS 0-3 at 90 days. This is likely due to similarly lower baseline NIHSS scores and a younger patient cohort, which may have contributed to improved recovery potential. Another important aspect of our study is the treatment time window, as the majority of our patients (65.4%) underwent EVT within six hours of symptom onset, a higher proportion than in other trials. This is particularly relevant as recent meta-analyses suggest that EVT effectiveness may decline in late-window patients, underscoring the need to explore whether NCCT-based selection remains valid beyond the conventional six-hour window.^12^

**Table 3.** Baseline Characteristics and Outcomes of Large Core Thrombectomy Trials and this study.

|  | Yoshimura et al.<br>RESCUE-JAPAN<br>LIMIT<br>n = 101 | A. Sarraj et al.<br>SELECT 2<br>n = 178 | Huo et al.<br>ANGEL-ASPECTS<br>n = 230 | Bendszus et al.<br>TENSION<br>n = 125 | V. Costalat et al.<br>LASTE<br>n = 159 | Zaidat et al.<br>TESLA<br>n = 152 | This Study<br>n = 52 |
| --- | --- | --- | --- | --- | --- | --- | --- |
| Study location | Japan | North America, Australia | China | Europe, Canada | Europe | US | Viet Nam |
| Median age (IQR) (years) | 76.6 (SD 10) | 66 (58-75) | 68 (61-73) | 73 (65-81) | 73 (66-79) | 65.5 (54-74) | 62,5 (58,5 - 71) |
| Sex: Male, n (%) | 55 (54.5) | 107 (60.1) | 135 (58.7) | 59 (55) | 82 (51.6) | 77 (50.7) | 30 (57.7) |
| Time from onset to randomization or Admit, Median (IQR), min | 229 (144-459)<br>(Random)<br>190 (85-390)<br>(Admit) | 544 (316-920)<br>NR | 453 (299-712)<br>338 (199-629) | 120 (72-210)<br>NR | 271 (199-351)<br>NR | 652.5 (333-942)<br>NR | 245 (107.5-469.5) |
| Time window<br>< 6h<br>≥ 6h | 71 (70.3)<br>30 (29.7) | NR | 82 (35.7)<br>148 (64.3) | 78(62.4)<br>47(37.6) | NR | 42 (27.6)<br>110 (72.4) | 35(67.3)<br>17(32.7) |
| ASPECST<br>Baseline<br>Median (IQR) | 3 (3-4) | 4 (3-5) | 3 (3-4) | 4 (3-5) | 2 (1-3) | 4 (3-5) | 4 (3 - 4) |
| NIHSS at Admit<br>Median (IQR) | 22 (18-26) | 19 (15-23) | 16 (13-20) | 19 (16-22) | 21 (18-24) | 19 (15-23) | 15 |
|  |  |  |  |  |  |  | (13-19.75) |
| IVT, n (%) | 27 (26.7) | 37 (20.8) | 66 (28.7) | 49 (39) | 55 (34.6) | 31 (20.4) | 16 (30.8) |
| mTICI 2b/3, n (%) | 86/100<br>(86) | 142<br>(79.8) | 183/226<br>(81) | 104<br>(83) | 130/151<br>(86.1) | 107/146<br>(73.3) | 41<br>(78.8) |
| mRS at 90 days<br>Median (IQR) | NR | 4 (3-6) | 4 (2-5) | 4 (3-6) | 4 (3-6) | 5 (3-6)<br>n=151 | 3.5 (3-5.5) |
| mRS 0-2 at 90 days, n (%) | 14<br>(14) | 36<br>(20.3) | 69<br>(30) | 21/124<br>(17) | 21/158<br>(13.3) | 22/151<br>(14.6) | 12<br>(23.1) |
| mRS 0-3 at 90 days, n (%) | 31<br>(31) | 67<br>(37.9) | 198<br>(47) | 39/124<br>(31) | 53/158<br>(33.5) | 45/151<br>(29.8) | 26<br>(50) |
| Mortality at 90 days, n (%) | 18<br>(18) | 68<br>(38.4) | 50<br>(21.7) | 49<br>(40) | 57/158<br>(36.1) | 53/150<br>(35.3) | 13<br>(25) |
| sICH (SITS-MOST), n (%) | 9 (9) | 1 (0.6) | 14 (6.1)<br>(HBC) | 7 (5)*<br>(HBC) | 5/157 (3.2)<br>15/157(9.6)<br>)HBC | 6/151<br>(4) | 5 (9.6)<br>13 (25) |
| Any intracranial hemorrhage, n (%) | 58<br>(58) | NR | 113<br>(49.1) | 68/124<br>(54.8) | 113/157<br>(72) | 86/148<br>(58.1) | 31<br>(59.6) |
| Decompression, n (%) | 10 (10) | NR | 17 (7.4) | 11 (9) | 14 (8.8) | 33 (21.9) | 5 (9.6) |

Abbreviations: ASPECTS (Alberta stroke programme early CT score), NIHSS (National Institutes of Health Stroke Scale), IVT (Intravenous thrombolysis), mTICI (The modified treatment in cerebral infarction), mRS (modified Rankin scale), sICH (symptomatic intracranial hemorrhage)

In terms of safety, this study reported an overall intracerebral hemorrhage rate of 59.6% and a symptomatic intracerebral hemorrhage (sICH) rate of 9.6%, comparable to other large clinical trials, particularly in Asia (RESCUE-JAPAN LIMIT: 9%, ANGEL-ASPECTS: 6.1%).^3,5^ This finding is crucial, as concerns about hemorrhagic risk have been a major barrier to expanding EVT in LIC patients. Additionally, our study found a 25% mortality rate, which was lower than in Western trials (SELECT 2, TENSION, LASTE, TESLA) and similar to Asian trials (RESCUE-JAPAN LIMIT, ANGEL-ASPECTS), indicating that NCCT-based selection is a safe approach and does not increase mortality risk. These findings suggest that simplified EVT protocols based on NCCT-CTA could be adopted in LMICs to expand access to thrombectomy, even for LIC patients.

Our study also highlighted that successful recanalization (mTICI ≥ 2b) was the strongest predictor of favorable outcomes. This finding is consistent with previous studies that emphasize the central role of achieving optimal reperfusion in determining EVT success, irrespective of infarct core size. A retrospective analysis of data from the German Stroke Registry also showed similar results, with 348 LIC patients, of whom 83.3% achieved mTICI 2b-3 recanalization. The successful reperfusion (mTICI 2b-3) group demonstrated better functional outcomes and lower mortality compared with the unsuccessful reperfusion group (mTICI 0-2a).^15^ A recently published multicenter retrospective aggregate cohort study on large ischemic volume thrombectomy further confirmed that the successful recanalization group had a significantly higher rate of favorable functional outcome (mRS 0-3) than the unsuccessful recanalization group (47.6% vs. 15.3%; OR, 5.02; 95% CI, 2.87– 8.76; P < 0.01).^16^ That study also demonstrated a significant reduction in mortality with successful recanalization, without an associated increase in the rate of symptomatic intracranial hemorrhage (sICH).^16^ These findings underscore the importance of achieving reperfusion to preserve salvageable tissue, which may play a more significant role in functional recovery than the initial infarct core volume. Collectively, they provide compelling evidence that successful recanalization is a key determinant of improved functional outcomes and reduced mortality, even in patients with large infarct cores.

Extended thrombectomy to the 24-hour window has been demonstrated in clinical trials since 2018.^17,18^ In recent years, a simple selection protocol using NCCT-CTA for thrombectomy in the late window (beyond 6 hours) has shown effectiveness in patients with small to medium-sized infarct cores (ASPECTS ≥6).^13,19^ Two NCCT-based clinical trials have explored extending the endovascular treatment window for patients with large infarct cores.^6,8^ The TENSION trial demonstrated efficacy in patients with ASPECTS 3-5 within 12 hours.^6^ Although the TESLA trial included ASPECTS 2-5 patients within 24 hours and did not meet its prespecified efficacy endpoint, it showed a strong trend.^8,20^ Subgroup analysis in our study suggests that thrombectomy in LIC patients may still be effective beyond 6 hours, as the ambulatory outcome in the late window group (41.2%) was comparable to that in the early window group, with no increased risk of sICH or death. However, the number of patients in the late window, particularly those treated 12 -24 hours after stroke onset, remained small. Future studies are needed to further expand the time window for large core patients.

Concerning ultra large core (ASPECTS 0-2), Our study indicated that EVT in ASPECTS 0-2 patients selected using NCCT was associated with poor outcomes. Therefore, expanding the treatment window for patients with extremely low ASPECTS (0-2) using a simple protocol appears unfeasible. These findings do not support EVT as a proven benefit for unlimited core volumes.

From the results of this study, we demonstrate the efficacy and safety of applying the NCCT-CTA protocol in treating patients with LICs. This is particularly meaningful for countries and centers with limited resources, as it helps expand EVT eligibility for more stroke patients, providing them with greater treatment opportunities. However, several limitations should be acknowledged. First, this was a single-center study, which may limit the generalizability of our findings to other hospitals with different stroke care infrastructures. Second, our sample size was relatively small (n=52), highlighting the need for larger multicenter studies to confirm these results, especially in special patient subgroups, such as the late-window patients (beyond 6 hours) or those with ultra-large infarct cores (ASPECTS 0–2). Third, our study lacked a control group of LIC patients who did not undergo EVT, preventing a direct comparison between EVT and medical management alone. Future comparative studies are needed to further validate the effectiveness and safety of EVT in LIC patients, particularly in resource-limited settings in LMICs.

## CONCLUSION

Our study provides real-world evidence that NCCT-CTA-based selection is a feasible, effective, and safe alternative to advanced imaging for EVT in large-core infarct patients, particularly in low-resource settings. The results demonstrate that NCCT can reliably guide EVT decision-making, yielding comparable functional outcomes and safety profiles to trials using advanced imaging modalities. Our findings also highlight that successful reperfusion (mTICI ≥2b) remains the strongest predictor of favorable outcomes, reinforcing its crucial role in improving functional recovery and reducing mortality, even in large infarct core patients selected by NCCT.

## Data Availability

The data that support the findings of this study are available from the corresponding author upon reasonable request.

## Acknowledgments

We sincerely acknowledge the staff of the Stroke Department and Da Nang Hospital for their dedication and invaluable support in patient care, treatment, and the execution of this study.

## Flow chart Patient Selection

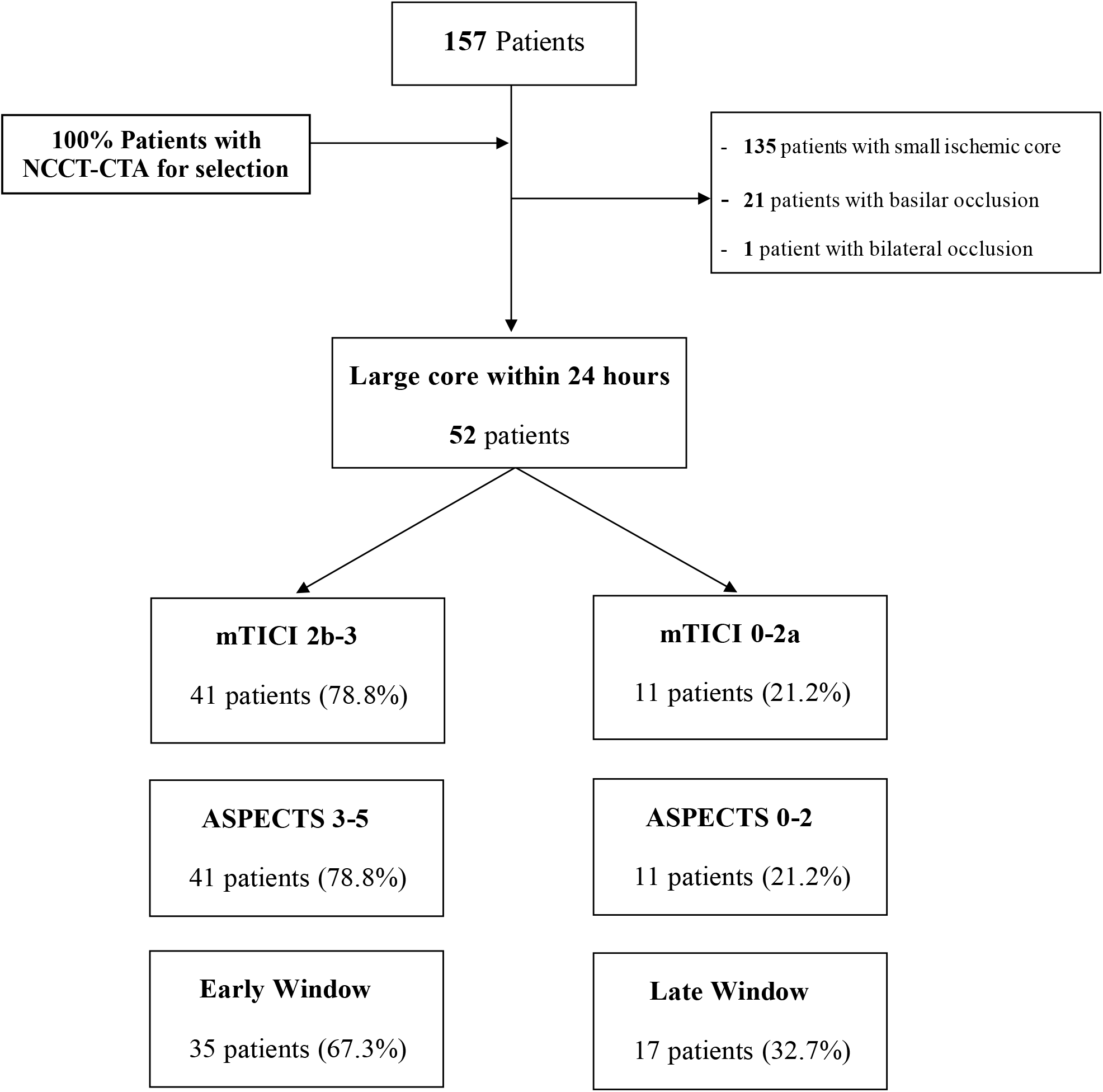

